# Cardiometabolic traits, sepsis and severe covid-19: a Mendelian randomization investigation

**DOI:** 10.1101/2020.06.18.20134676

**Authors:** Mark J Ponsford, Apostolos Gkatzionis, Venexia M Walker, Andrew J Grant, Robyn E Wootton, Luke S P Moore, Segun Fatumo, Amy M Mason, Verena Zuber, Cristen Willer, Humaira Rasheed, Ben Brumpton, Kristian Hveem, Jan Kristian Damås, Neil Davies, Bjørn Olav Åsvold, Erik Solligård, Simon Jones, Stephen Burgess, Tormod Rogne, Dipender Gill

**Affiliations:** Immunodeficiency Centre or Wales, University Hospital Wales, Heath Park, Cardiff, UK; Division of Immunology, Infection, and Inflammation, Tenovus Institute, Cardiff University, Cardiff, UK; MRC Biostatistics Unit, School of Clinical Medicine, University of Cambridge, Cambridge, UK; MRC Integrative Epidemiology Unit, Bristol Medical School, University of Bristol, Bristol, UK; Department of Surgery, University of Pennsylvania Perelman School of Medicine, Philadelphia, USA; National Institute for Health Research Health Protection Research Unit in Healthcare Associated Infections and Antimicrobial Resistance, Imperial College London, UK; Chelsea & Westminster NHS Foundation Trust, London, UK; Imperial Biomedical Research Centre, Imperial College London and Imperial College NHS Healthcare Trust, London, UK; Department of Non-Communicable Diseases Epidemiology, London School of Hygiene & Tropical Medicine, UK; Cardiovascular Epidemiology Unit, Department of Public Health and Primary Care, University of Cambridge, Cambridge, UK; National Institute for Health Research Cambridge Biomedical Research Centre, University of Cambridge and Cambridge University Hospitals, Cambridge, UK; Department of Epidemiology and Biostatistics, School of Public Health, Imperial College London, London, UK; Departments of Internal Medicine, Human Genetics and Computational Medicine & Bioinformatics, University of Michigan, Ann Arbor, MI, USA; K.G. Jebsen Center for Genetic Epidemiology, Department of Public Health and Nursing, Norwegian University of Science and Technology, NTNU, Trondheim, Norway; Department of Thoracic Medicine, St. Olavs Hospital, Trondheim University Hospital, Trondheim, Norway; Centre of Molecular Inflammation Research, Department of Clinical and Molecular Medicine, Norwegian University of Science and Technology, NTNU, Trondheim, Norway; Department of Infectious Diseases, St Olavs Hospital, Trondheim University Hospital, Trondheim, Norway; Gemini Center for Sepsis Research, Department of Circulation and Medical Imaging, Norwegian University of Science and Technology, NTNU, Trondheim, Norway; Department of Epidemiology and Biostatistics, Medical School Building, St Mary’s Hospital, Imperial College London, London, UK

## Abstract

**Objectives:** To investigate whether there is a causal effect of cardiometabolic traits on risk of sepsis and severe covid-19.

**Design:** Mendelian randomisation analysis.

**Setting:** UK Biobank and HUNT study population-based cohorts for risk of sepsis, and genome-wide association study summary data for risk of severe covid-19 with respiratory failure.

**Participants:** 12,455 sepsis cases (519,885 controls) and 1,610 severe covid-19 with respiratory failure cases (2,205 controls).

**Exposure:** Genetic variants that proxy body mass index (BMI), lipid traits, systolic blood pressure, lifetime smoking score, and type 2 diabetes liability - derived from studies considering between 188,577 to 898,130 participants.

**Main outcome measures:** Risk of sepsis and severe covid-19 with respiratory failure.

**Results:** Higher genetically proxied BMI and lifetime smoking score were associated with increased risk of sepsis in both UK Biobank (BMI: odds ratio 1.38 per standard deviation increase, 95% confidence interval [CI] 1.27 to 1.51; smoking: odds ratio 2.81 per standard deviation increase, 95% CI 2.09-3.79) and HUNT (BMI: 1.41, 95% CI 1.18 to 1.69; smoking: 1.93, 95% CI 1.02-3.64). Higher genetically proxied BMI and lifetime smoking score were also associated with increased risk of severe covid-19, although with wider confidence intervals (BMI: 1.75, 95% CI 1.20 to 2.57; smoking: 3.94, 95% CI 1.13 to 13.75). There was limited evidence to support associations of genetically proxied lipid traits, systolic blood pressure or type 2 diabetes liability with risk of sepsis or severe covid-19. Similar findings were generally obtained when using Mendelian randomization methods that are more robust to the inclusion of pleiotropic variants, although the precision of estimates was reduced.

**Conclusions:** Our findings support a causal effect of elevated BMI and smoking on risk of sepsis and severe covid-19. Clinical and public health interventions targeting obesity and smoking are likely to reduce sepsis and covid-19 related morbidity, along with the plethora of other health-related outcomes that these traits adversely affect.

**Summary boxes:** *What is already known on this topic:*

- Sepsis and severe covid-19 are major contributors to global morbidity and mortality.
- Cardiometabolic risk factors have been associated with risk of sepsis and severe covid-19, but it is unclear if they are having causal effects.

*What this study adds:*

- Using Mendelian randomization analyses, this study provides evidence to support that higher body mass index and lifetime smoking score both increase risk of sepsis and severe covid-19 with respiratory failure.
- Clinical and public health interventions targeting obesity and smoking are likely to reduce sepsis and covid-19 related morbidity, along with the plethora of other health-related outcomes that these traits adversely affect.

## Introduction

Sepsis is defined as life-threatening organ dysfunction and is characterised by a dysregulated host inflammatory and immune response to infection.^1^ It is a major contributor to the global burden of disease and in 2017 alone almost 49 million sepsis cases were estimated to have occurred, causing 20% of all deaths globally.^2^ Despite implementation of protocolised pathways seeking to improve care quality and individualise management through goal-directed therapy, sepsis remains a leading cause of critical illness in developed countries.^3 4^ A greater understanding of host factors that predispose to sepsis is urgently needed to improve primary prevention and support the development of mechanism-guided treatment. The severe acute respiratory coronavirus 2 (SARS-COV-2) causing coronavirus disease 2019 (covid -19) emerged in late 2019, quickly developing into a pandemic.^5^ Among patients hospitalised with covid-19, many develop sepsis.^6^ As with sepsis, it is essential to identify risk factors for severe covid-19 in order to reduce the global burden of disease.

Several observational studies have supported an association between obesity and infection.^7-11^ In the largest reported to date, Winter-Jensen *et al*. followed 101,447 individuals from the Copenhagen General Population Study cohort over 8.8 years.^9^ Here, elevated body mass index (BMI) was associated with increased risk of sepsis, urinary tract, and skin infection.^9^ Epidemiological evidence has also identified an association of obesity with increased risk of severe covid-19 in the current pandemic.^12- 16^ However, causality cannot be reliably ascertained from observational studies alone, due to issues such as confounding and reverse causation.^17^ For example, the phenotype of obesity is entangled with a range of wider cardiometabolic traits such as smoking, altered lipid profile, and diabetes, which represent potential confounding exposures.^18^

In the Mendelian randomization (MR) approach, genetic variants are used to proxy an exposure in the investigation of its effect on an outcome. The use of genetic variants that are randomly allocated at conception means that MR is less vulnerable to the confounding and reverse causation bias that can limit causal inference when using observational research approaches.^19^ The opportunity now exists to perform genetic association analyses of sepsis in large-scale population cohorts, and leverage these for MR.^9 20 21^ For example, a recent MR study of UK Biobank participants supported that elevated high-density lipoprotein cholesterol (HDL-C) confers a protective effect on sepsis risk.^21^ Similarly, genome-wide association summary data for severe covid-19 with respiratory failure have recently been made publicly available.^22^ However, to our knowledge no MR studies have systematically investigated the association between a range of cardiometabolic traits and risk of sepsis or severe covid-19.

Here, we use data from large-scale genetic association studies to identify genetic proxies for BMI, lifetime smoking, lipid traits, systolic blood pressure (SBP) and type 2 diabetes mellitus (T2DM), and apply these in MR analyses investigating risk of sepsis in the UK Biobank (UK) and the HUNT study (Norway). We further extend our analyses to investigate the outcome of severe covid-19 with respiratory failure. Our findings offer insight into the causal effects of cardiometabolic traits on risk of sepsis and severe covid-19, and therefore identify modifiable targets amenable to clinical and public health intervention.

## Methods

### Genetic association estimates

#### Cardiometabolic traits

We investigated the association of genetically proxied levels of five cardiometabolic traits with sepsis risk using univariable MR analysis: BMI, lifetime smoking (referred to hereafter as smoking), low-density lipoprotein cholesterol (LDL-C), SBP, and liability to T2DM. Genetic association estimates were obtained from the publicly available GWAS summary data sources listed in **Table 1**. Genetic variants selected as instruments were uncorrelated (r^2^ < 0.001) single-nucleotide polymorphisms (SNPs) associated with the corresponding trait at a genome-wide level of statistical significance (p < 5×10^−8^), with clumping performed using the TwoSampleMR package in R.^23^ Given the previously identified association between HDL-C and risk of infectious disease,^21^ and the high genetic correlation between circulating LDL-C, HDL-C and triglycerides,^24^ we also performed multivariable MR analysis investigating the direct associations of genetically predicted levels of each of these traits on sepsis risk. Genetic association estimates for HDL-C and triglycerides were obtained from the same study as for LDL-C.^24^ Correlated variants for multivariable MR were clumped based on the lowest p-value for their association with any of the three lipid traits.^23^

**Table 1.**
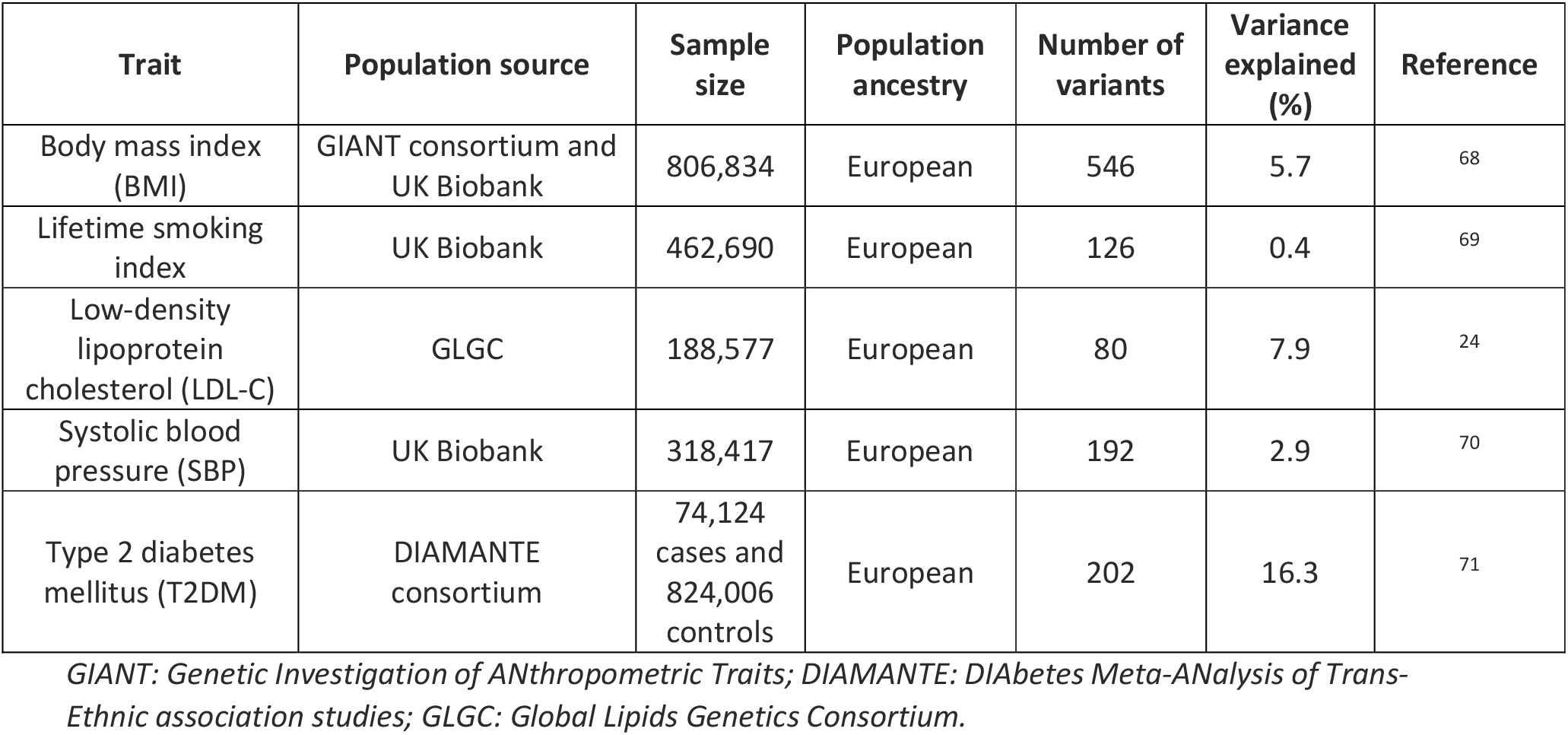
Exposure trait genetic summary data sources.

#### Risk of sepsis

##### UK Biobank

We generated GWAS summary data for sepsis in the UK Biobank, a national health research resource of 502,647 participants aged 40-69 years. Participants were recruited from assessment centres across the UK between 2006 and 2010.^25 26^ We defined sepsis using a previously published list of explicit ICD-9 and ICD-10 codes derived by a panel of experts in critical care, infectious diseases, paediatrics, and sepsis epidemiology (**Supplementary Table 1)**.^2^ Briefly, this was a binary variable based on the presence of one or more codes as a main or secondary diagnosis in the hospital inpatient admissions data between 1996 and 31^st^ March 2017 (UK Biobank data-fields 41270 and 41271) or as a primary or secondary cause of death in the death registry data between April 2006 and 31^st^ March 2020 (UK Biobank data-fields 40001 and 40002). Data were analysed using the Medical Research Council Integrative Epidemiology Unit GWAS pipeline.^27^ A BOLT-LMM linear mixed model was used to account for relatedness between individuals and population stratification, with adjustment for age, sex and genotyping chip. Exclusions were made for individuals whose genetic sex did not match their reported sex, individuals with sex chromosome karyotypes putatively different from XX or XY, individuals who were outliers in heterozygosity and missing rates, and individuals with high levels of relatedness (3rd degree or greater) to more than 200 other individuals. Genetic association estimates were converted from an absolute risk difference scale to log odds ratios prior to MR analysis.

##### HUNT study

The population-based HUNT study collected information on 126,000 individuals between 1984 and 2019 from inhabitants of the Norwegian county of Trøndelag. Data collection was conducted in 4 survey waves, between 1984 to 2019, of which data from HUNT2 (1995-1997) and HUNT3 (2006-2008) on subjects 20 years and older were used in the current work.^28^ Genotyping and quality control for the HUNT study was performed as previously described.^29^ Sepsis was defined the same way in the HUNT Study as in UK Biobank. Association tests were carried out using SAIGE, a generalized logistic mixed model that accounts for kinship and imbalance in the number of cases and controls.^30^ We adjusted for age, sex, genotype batch, and the five first ancestry-informative principal components. The principal components were estimated with the TRACE software package, with 938 individuals from the Human Genome Diversity Project serving as reference.^31 32^

#### Risk of severe covid-19 with respiratory failure

Summary genetic association estimates for risk of severe covid-19 with respiratory failure were obtained from a genome-wide association study performed in 1610 cases and 2205 controls (with no or mild covid-19 symptoms) in Italy and Spain.^22^ All cases had a positive SARS-CoV-2 viral RNA polymerase-chain-reaction test from nasopharyngeal swabs or other biologic fluids, and respiratory failure defined as the use of oxygen supplementation or mechanical ventilation at any point during their hospital admission.^22^ Cohorts details have been described previously, and data from the GWAS that adjusted for the first ten ancestry-informative principal components, age and sex were used in our analyses.^22^

### Mendelian randomization analysis

The main univariable analyses estimating the association of genetically proxied levels of each exposure with risk of sepsis and severe covid-19 with respiratory failure were performed using the inverse-variance weighted (IVW) method and a random-effects model.^33^ The IVW method combines the causal effect estimates from each individual genetic variant (computed as the ratio of the variant-sepsis association to the variant-exposure association). It provides a statistically consistent estimator of the true causal effect as long as all genetic variants are valid instrumental variables.^34^ If any of the genetic variants are invalid instruments due to pleiotropy (where the variant affects sepsis or severe covid-19 risk through pathways unrelated to the exposure), the estimator is still statistically consistent as long as the pleiotropy is balanced (that is, the average of the pleiotropic effects of each genetic variant on the outcome are equal to zero).

Sensitivity analyses for univariable MR were performed using the weighted median method,^35^ a weighted mode-based method,^36^ the contamination mixture method,^37^ and the MR-Egger method.^38^ Each of these methods provides a statistically consistent estimator of the true causal effect under different sets of assumptions. The weighted median, mode-based and contamination mixture methods all allow some genetic variants to be invalid instruments and are robust to outliers in the genetic variant-specific causal effect estimates. The MR-Egger method allows all genetic variants to be invalid but requires the pleiotropic effects that relate to sepsis or severe covid-19 risk through pleiotropic pathways to be uncorrelated with the genetic variant-exposure associations, and is also sensitive to outliers. The intercept test of the MR-Egger method was used to test for unbalanced, or directional, pleiotropy. The lower the p-value from the intercept test, the greater the evidence to support the presence of directional pleiotropy.

To estimate the direct effect of genetically predicted LDL-C, HDL-C and triglycerides on risk of sepsis and severe covid-19, summary data multivariable MR was performed.^39-41^ Here, the variant-outcome genetic association estimates were regressed on the three variant-lipid trait estimates, weighted for the precision of the variant-outcome risk association, and with the intercept fixed to zero.^41^

Causal effect estimates are expressed as odds ratios per standard deviation increase in genetically predicted levels of the exposure for continuous traits, and per unit increase in the log odds ratio of the exposure for binary traits. All analyses were performed using the MendelianRandomization package in R.^42^

### Ethical approval, data availability and reporting

The data used in this work obtained relevant participant consent and ethical approval. All variants used as instruments and their genetic association estimates were selected from publicly available data sources, and are provided in **Supplementary Tables 2-6**. The results from the analyses performed in this work are presented in the main manuscript or its supplementary files. This paper has been reported based on recommendations by the STROBE-MR Guidelines (**Supplementary Research Checklist**).^43^ The study protocol and details were not pre-registered. Code for analyses are available via the online Github repository (https://github.com/agkatzionis/Sepsis-MR).

**Table 2.** UK Biobank and HUNT cohort descriptions.

|  | UK Biobank |  | HUNT |  |
| --- | --- | --- | --- | --- |
|  | All considered participants | Cases | All considered participants | Cases |
| Number | 462,918 | 10,154 | 69,422 | 2,301 |
| Female sex, (%) | 251,078<br>(54%) | 5,655<br>(56%) | 36,784<br>(53.0%) | 1,136<br>(49.4%) |
| Median age at recruitment, years (interquartile range) | 58<br>(50 to 63) | 60<br>(52 to 65) | 46<br>(34 to 62) | 63<br>(50 to 71) |
| Ever smoker, (%) | 282,402<br>(61%) | 6,715<br>(66%) | 38,591<br>(57%) | 1,495<br>(66.6%) |
| Mean body mass index, (kg/m<sup>2</sup>, SD) | 27.4<br>(4.8) | 28.2<br>(5.6) | 26.4<br>(4.1) | 27.4<br>(4.5) |
| Mean low-density lipoprotein cholesterol, (mmol per litre, SD) | 3.6<br>(0.9) | 3.4<br>(0.9) | 3.6<br>(1.1) | 3.9<br>(1.1) |
| Mean systolic blood pressure, (mmHg, SD) | 135.9<br>(18.6) | 136.1<br>(19.4) | 135.1<br>(21.0) | 143.6<br>(23.5) |
| Type 2 diabetes mellitus, (%) | 24,903<br>(5%) | 1499<br>(15%) | 2,130<br>(3%) | 173<br>(8%) |

*SD: standard deviation.*

### Patient and public involvement

No patients or participants were involved in setting the research question or the outcome measures, nor were they involved in developing plans for design or implementation of the study. No patients were asked to advise on interpretation or writing up of results. The published manuscript relating to this work will be sent to the UK Biobank resource for appropriate dissemination to participants.

## Results

### UK Biobank and HUNT cohort descriptions

The baseline characteristics of the participants used to obtain sepsis genetic association estimates are shown in **Table 2**. The considered UK Biobank sample consisted of 462,918 individuals of European ancestry who had phenotype data and passed genotype inclusion criteria (54% female; median age at recruitment = 58 years; interquartile range = 50 to 63 years). Overall, 61% of the sample had ever smoked, with a mean BMI of 27.4 kg/m^2^, LDL-C of 3.6 mmol/L, SBP of 136 mm Hg, and a 5% prevalence of T2DM (**Table 2**). The HUNT sample considered 69,422 participants. These individuals were younger at study entry than those in the UK Biobank study (median age 46 years versus 58 years) with similar sex ratio (53% female). A greater proportion of participants in UK Biobank had a lifetime history of smoking (61% vs 57%), and type 2 diabetes (5% versus 3%). Distributions of BMI, LDL-C, and SBP were similar in both UK Biobank and HUNT participants. Applying the explicit definition of sepsis used by Rudd et al,^2^ we identified 10,154 cases within UK Biobank, of which 56% were female and 66% smokers. Mean BMI was 28.2 kg/m^2^, greater than 27.4 kg/m^2^ in the control group of 452,764 individuals. Within HUNT, we identified 2,301 cases of sepsis and 67,121 controls. Cases tended to be older than controls, with a median age at recruitment of 63 years compared to 46 years.

### Associations of genetically-proxied cardiometabolic traits with risk of sepsis and severe covid-19

In univariable MR analysis of the UK Biobank cohort, higher genetically proxied BMI and lifetime smoking score were associated with increased risk of developing sepsis (**Figure 1**). For every standard deviation (4.8 kg/m^2^) increase in genetically proxied BMI, the odds ratio (OR) for developing sepsis was 1.38 (95% confidence interval: 1.27 to 1.51). For every standard deviation in lifetime smoking score, the OR for developing sepsis was 2.81 (95% confidence interval: 2.09 to 3.79). Similar evidence for an effect of BMI and smoking was also found in MR analyses considering sepsis in the HUNT study (BMI: OR 1.41, 95% confidence interval: 1.18 to 1.69; smoking: OR 1.93, 95% confidence interval: 1.02 to 3.64) although estimates had wider confidence intervals. Higher genetically proxied BMI and smoking were also associated with increased risk of severe covid-19 with respiratory failure (BMI: OR 1.75, 95% confidence interval: 1.20 to 2.57; smoking: OR 3.94, 95% confidence interval 1.13 to 13.75) (**Figure 2**). There was limited evidence for effects of LDL-C, SBP, or T2DM on risk of sepsis (in either UK Biobank or HUNT) or severe covid-19 with respiratory failure.

**Figure 1:**
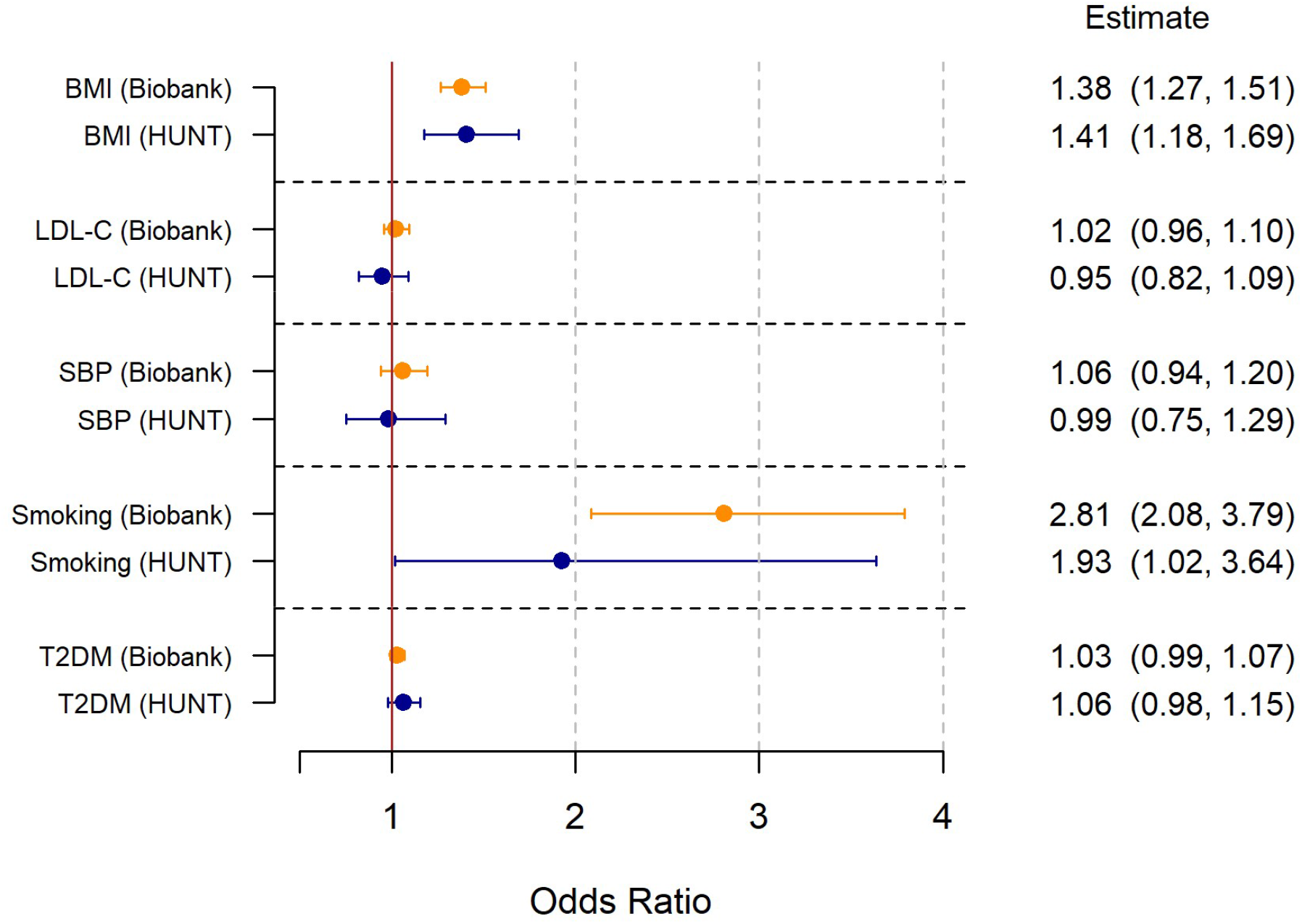
Results of the univariable Mendelian randomization analysis investigating the association of genetically proxied cardiometabolic traits with risk of sepsis.Forest plot showing inverse-variance weighted Mendelian randomization estimates for the association between cardiometabolic traits and risk of sepsis in the UK Biobank (n = 462,918) and HUNT cohorts (n = 69,422). Results are expressed per standard deviation increase in genetically proxied levels of the exposure for continuous traits (BMI, LDL, SBP, and smoking), and per unit increase in log odds ratio for genetically proxied T2DM liability. BMI: body mass index; LDL-C: low-density lipoprotein cholesterol; SBP: systolic blood pressure; T2DM: type 2 diabetes mellitus.

**Figure 2:**
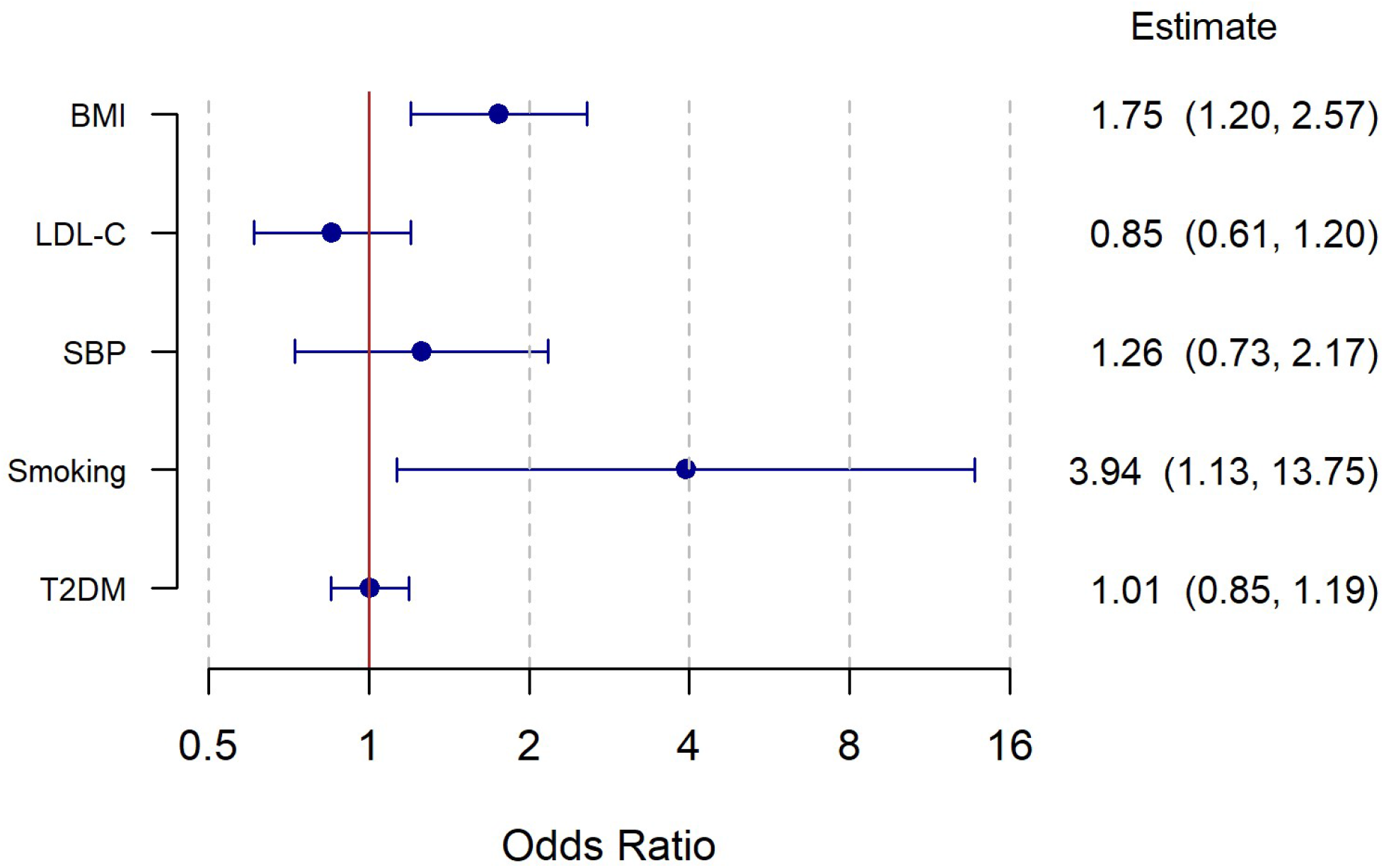
Results of the univariable Mendelian randomization analysis investigating the association of genetically proxied cardiometabolic traits with risk of severe covid-19 with respiratory failure. Forest plot showing inverse-variance weighted Mendelian randomization estimates for the association between cardiometabolic traits and risk of severe covid-19 with respiratory failure. Results are expressed per standard deviation increase in genetically proxied levels of the exposure for continuous traits (BMI, LDL, SBP, and smoking), and per unit increase in log odds ratio for genetically proxied T2DM liability. BMI: body mass index; LDL-C: low-density lipoprotein cholesterol; SBP: systolic blood pressure; T2DM: type 2 diabetes mellitus. A logarithmic scale is used on the x-axis.

### Sensitivity analyses

To investigate the robustness of our findings, we employed multiple MR methods making distinct assumptions about the inclusion of pleiotropic variants. The association of genetically proxied BMI with risk of sepsis was consistent when performing MR-Egger, weighted median, and contamination mixture within UK Biobank (**Supplementary Figure 1**), with a similar point estimate but 95% confidence intervals that included the null when using the mode-based estimator. Findings for smoking exposure were consistent across all methods, but produced 95% confidence intervals that crossed the null when using MR-Egger. Similar findings for associations of genetically proxied BMI and smoking with risk of sepsis were observed across all methods in the smaller HUNT cohort (**Supplementary Figure 1**), with wider 95% confidence intervals.

In the analyses of severe covid-19 with respiratory failure, the MR sensitivity analyses considering genetically proxied BMI as the exposure produced similar estimates to the main analysis (**Supplementary Figure 2**), and only the 95% confidence intervals for MR-Egger crossed the null. The MR sensitivity analyses considering genetically proxied lifetime smoking score as the exposure also produced similar estimates to the main analysis, with the exception of MR-Egger, which had very wide confidence intervals (**Supplementary Figure 2**). The 95% confidence interval for the MR-Egger intercept term included the null in analyses for all the considered exposures and outcomes (**Supplementary table 7**).

To investigate confounding related to genetic correlation between the variants used to proxy lipid traits, we performed multivariable MR analysis. Lower levels of genetically-proxied HDL-C had a point estimate that indicated lower risk of developing sepsis within the UK Biobank, however the 95% confidence intervals included the null (**Figure 3A**), and this was not observed within HUNT (**Figure 3B**). Overall, there was little evidence to support an association of the considered lipid traits (LDL-C, HDL-C, and triglycerides) with sepsis in these analyses. Similarly, there was little evidence supporting an association of the considered lipid traits with risk of severe covid-19 with respiratory failure (**Figure 4**).

**Figure 3:**
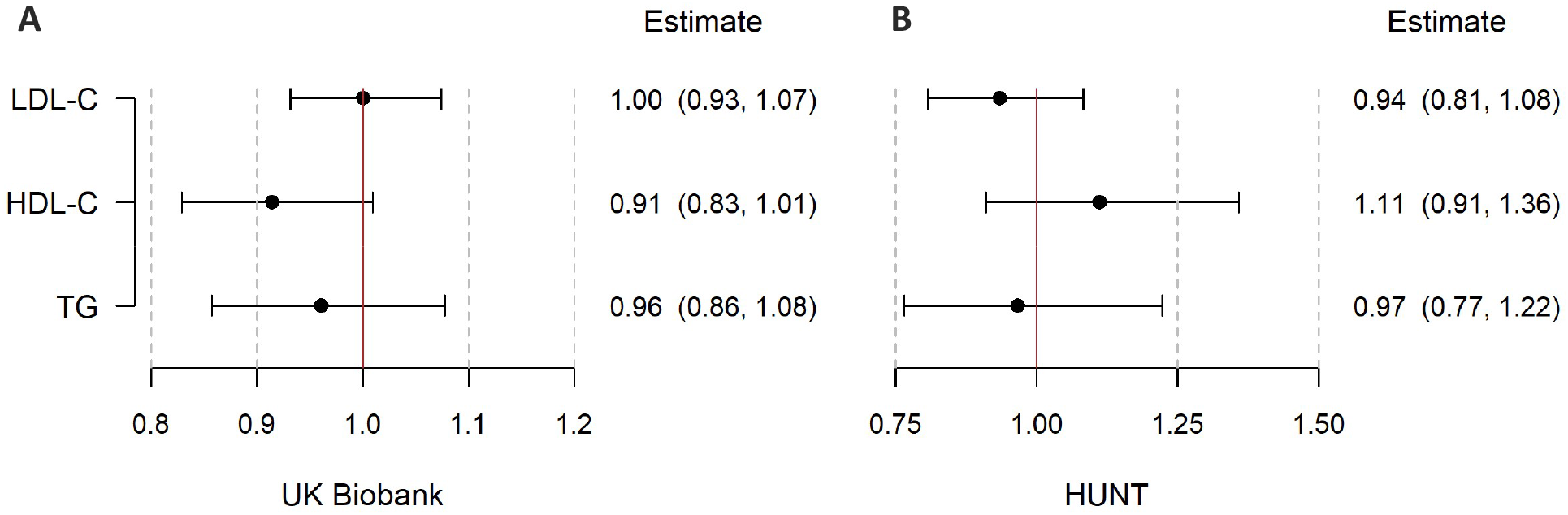
Multivariable Mendelian randomization analysis for association of genetically proxied lipid traits with risk of sepsis in UK Biobank and HUNT. Forest plot showing multivariable Mendelian randomization analyses for the association between genetically proxied lipid traits and risk of sepsis in A: UK Biobank (n = 462,918) and B: HUNT cohorts (n = 69,422). Odds ratio and 95% confidence interval are expressed for a one standard deviation increase in genetically proxied levels of the lipid trait. LDL-C: low-density lipoprotein cholesterol, HDL-C: high-density lipoprotein cholesterol; TG: triglycerides.

**Figure 3:**
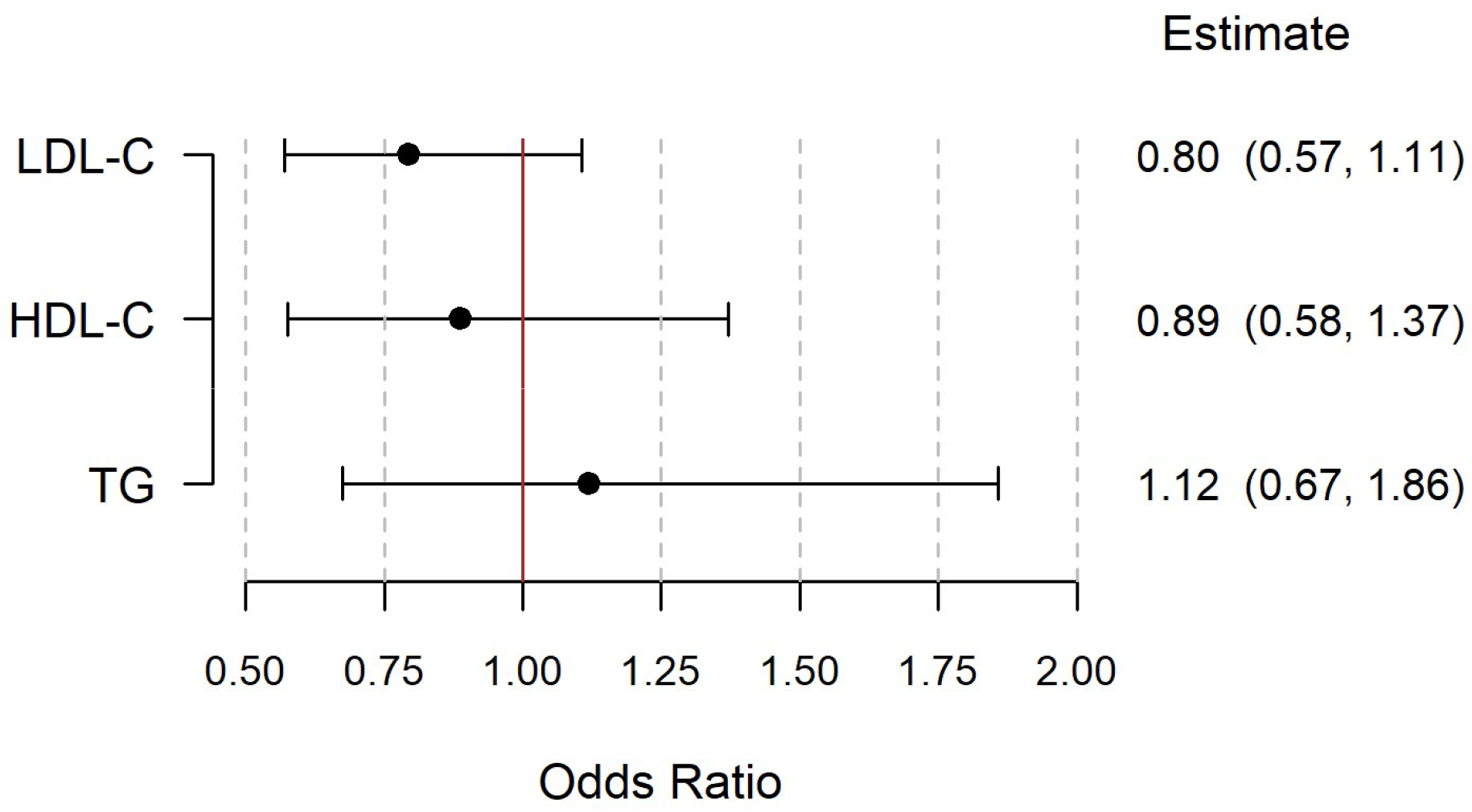
Multivariable Mendelian randomization analysis for association of genetically proxied lipid traits with risk of severe sepsis with respiratory failure. Forrest plot showing multivariable Mendelian randomization analyses for the association between genetically proxied lipid traits and risk of severe covid-19 with respiratory failure. Odds ratio and 95% confidence interval are expressed for a one standard deviation increase in genetically proxied levels of the lipid trait. LDL-C: low-density lipoprotein cholesterol, HDL-C: high-density lipoprotein cholesterol; TG: triglycerides.

## Discussion

In this MR study, we used publicly-available large-scale GWAS data to identify genetic variants that proxy the effect of modifying cardiometabolic traits, and investigated their association with risk of sepsis and severe covid-19 with respiratory failure. We found that higher genetically proxied BMI and smoking were associated with increased risk of sepsis in both the UK Biobank and HUNT study, and also with increased risk of severe covid-19. Genetically proxied lipid traits, SBP and T2DM liability had limited evidence for an association with risk of sepsis or severe covid-19, and concordant results were obtained in sensitivity analysis using MR methods that make different assumptions about the inclusion of pleiotropic variants. Taken together, our findings support the hypothesis that elevated BMI and smoking result in increased susceptibility to sepsis and severe covid-19.

A range of potential mechanisms have been suggested to link obesity and sepsis, including obesity-driven immune system dysregulation,^8 17^ related co-morbidities,^18^ and respiratory dysfunction.^44^ Smoking is similarly a well-recognised driver of systemic immune dysregulation.^45 46^ An exaggerated inflammatory response appears a major driver of immunopathology in SARS-CoV-2 infected patients,^47-49^ with early reports supporting benefits of targeting the acute inflammatory response in severe covid-19 cases.^50-52^ In contrast, emerging evidence suggests that diabetic patients show a similar inflammatory and lipid responses to non-diabetics during sepsis, with similar rates of sepsis-related mortality to the general population. ^53^

Obesity remains a leading, preventable threat to global health, for which the incidence continues to rise at an alarming rate.^54^ A recent report estimated that by 2030, almost half of adults in the US will have a BMI greater than 30 kg/m^2^.^55^ The prevalence of morbid obesity (BMI ≥ 40kg/m^2^) is projected to approach 11% within the UK by 2035, with population sub-groups exceeding 20%.^56^ Obesity has been shown to be relevant to infection risk in previous epidemiological studies,^9 10^ however this is the first MR analysis supporting a causal relationship. To our knowledge, this is also the first MR analysis exploring the relationship between smoking and sepsis risk. Our findings extend observational epidemiological studies that describe a dose-response relationship between tobacco smoke exposure and risk of pneumonia.^11 57^ These found active smokers to be over a third more likely to develop an influenza-like illness than non-smokers.^58^ According to the 2019 World Health Organization report on the global tobacco epidemic, smoking prevalence remains high globally at around 20%. Whilst the estimated prevalence of daily smoking has fallen since 1980, population growth has led to a net increase in the number of smokers.^59^ In 2018, the UK adult smoking prevalence remained at 15%,^60^ despite a range of public health measures aimed at reducing this.^60^ This figure is likely to mask considerable heterogeneity within clinical, demographic, and socioeconomic groups. For instance, persons with mental illness are about twice as likely to smoke as other persons, ^61^ have a lower quit rate,^62^ and elevated rates of obesity.^63^ This convergence of risk factors is consistent with the reported elevation in infection rate within this patient group.^64^

Our study has a number of strengths. The employed MR framework is less susceptible to the environmental confounding and reverse causation that can limit causal inference from being drawn in observational studies. We performed a range of MR methods that vary in their requisite assumptions regarding the inclusion of pleiotropic variants. The similar findings for genetically proxied BMI and smoking across these methods, although with expected variation in precision,^65^ support the robustness of our findings. Similarly, the association of genetically proxied BMI and smoking with risk of sepsis was observed in both the UK Biobank and the HUNT study. To our knowledge, this current work represents the largest genetic analysis of sepsis published to date. By using an explicit definition of sepsis that did not focus on any particular type of infection,^2^ our findings are more likely to be generalizable across different causes of sepsis. We formally tested this hypothesis by extending MR analyses to consider risk of severe covid-19, which produced results convergent with observational data.^12 13^ Genetic associations with covid-19 diagnosis are likely to be influenced by selection bias, as diagnosis is not possible without a positive test, and community testing is more common in certain professions and socio-economic groups. However, genetic associations with severe covid-19 are less likely to be subject to selection because all those with severe symptoms will likely seek medical treatment and hence be subject to testing.

Our study also has limitations, including that the findings currently lack external validity. Specifically, our investigation was conducted in European ancestry participants, and the findings may not necessarily apply to other ethnic groups. It is also important to appreciate that this MR study considers the lifelong effect of genetic modification of cardiometabolic traits, which is not the same as a clinical or public health intervention to modify them. The MR effect estimates should therefore not be directly extrapolated for this purpose, but should rather be used as evidence to support a causal relationship. Although we explored the association of genetically proxied T2DM liability with risk of sepsis, we were not able to explore genetically proxied T2DM itself, as this is a binary trait.^66^ Thus, there may be a causal relationship between diabetes (or glycaemic control) with sepsis risk that our study design is unable to uncover. Indeed, in a retrospective cohort study of the UK Clinical Practice Research Database, patients with evidence of poor glycaemic control (with mean HbA_1c_ ≥ 11%, or 96.7mmol/mol) were almost three times as likely to be hospitalized with infection as age- and sex-matched non-diabetics.^67^ Finally, our study also had limited statistical power, as apparent from the confidence intervals of the result. Larger genetic analyses of sepsis and severe covid-19 in the future may offer the opportunity to improve on this.

In conclusion, we leveraged large-scale genetic summary data to investigate the effect of cardiometabolic traits on risk of sepsis and severe covid-19 using robust MR analysis methods in a range of distinct populations. Our findings support a causal effect of elevated BMI and smoking on susceptibility to sepsis and severe covid-19, and add to the extensive body of evidence already supporting detrimental consequences of these traits across a range of health outcomes. These findings advocate expanding clinical and public health interventions to combat the global threats of obesity and smoking, particularly in the current covid-19 pandemic.

## Data Availability

All variants used as instruments and their genetic association estimates were selected from publicly available data sources, and are provided in Supplementary Tables 2-6. The results from the analyses performed in this work are presented in the main manuscript or its supplementary files. This paper has been reported based on recommendations by the STROBE-MR Guidelines (Supplementary Research Checklist).41 The study protocol and details were not pre-registered. Code for analyses are available via the online Github repository (https://github.com/agkatzionis/Sepsis-MR).

## Acknowledgements

This research was conducted using the UK Biobank Resource under Application Number 743915825. Quality Control filtering of the UK Biobank data was conducted by R. Mitchell, G. Hemani, T. Dudding, Corbin, S. Harrison, L. Paternoster as described in the published protocol (doi: 10.5523/bris.1ovaau5sxunp2cv8rcy88688v). The MRC IEU UK Biobank GWAS pipeline was developed by B. Elsworth, R. Mitchell, C. Raistrick, L. Paternoster, G. Hemani, T. Gaunt (doi: 10.5523/bris.pnoat8cxo0u52p6ynfaekeigi). The views expressed are those of the authors and not necessarily those of the National Health Service, National Institute for Health Research, or Department of Health and Social Care. The HUNT Study is a collaboration between the Nord-Trøndelag Health (HUNT) Research Centre (Faculty of Medicine and Health Sciences, NTNU, Norwegian University of Science and Technology), Nord-Trøndelag County Council, Central Norway Regional Health Authority, and the Norwegian Institute of Public Health. The genotyping in HUNT was financed by the National Institutes of Health; University of Michigan; the Research Council of Norway; the Liaison Committee for Education, Research and Innovation in Central Norway; and the Joint Research Committee between St Olavs Hospital and the Faculty of Medicine and Health Sciences, NTNU.

## References

1. Singer M, Deutschman CS, Seymour CW, et al. The Third International Consensus Definitions for Sepsis and Septic Shock (Sepsis-3). JAMA 2016;315(8):801–10. doi: 10.1001/jama.2016.0287

2. Rudd KE, Johnson SC, Agesa KM, et al. Global, regional, and national sepsis incidence and mortality, 1990–2017: analysis for the Global Burden of Disease Study. The Lancet 2020;395(10219):200–11. doi: https://doi.org/10.1016/S0140-6736(19)32989-7

3. Dombrovskiy VY, Martin AA, Sunderram J, et al. Rapid increase in hospitalization and mortality rates for severe sepsis in the United States: A trend analysis from 1993 to 2003*. Critical Care Medicine 2007;35(5)

4. Bouza C, López-Cuadrado T, Saz-Parkinson Z, et al. Epidemiology and recent trends of severe sepsis in Spain: a nationwide population-based analysis (2006-2011). BMC Infectious Diseases 2014;14(1):3863. doi: 10.1186/s12879-014-0717-7

5. Zhu N, Zhang D, Wang W, et al. A Novel Coronavirus from Patients with Pneumonia in China, 2019. N Engl J Med 2020;382(8):727–33. doi: 10.1056/NEJMoa2001017

6. Zhou F, Yu T, Du R, et al. Clinical course and risk factors for mortality of adult inpatients with COVID-19 in Wuhan, China: a retrospective cohort study. Lancet 2020;395(10229):1054–62. doi: 10.1016/S0140-6736(20)30566-3

7. Ghilotti F, Bellocco R, Ye W, et al. Obesity and risk of infections: results from men and women in the Swedish National March Cohort. International Journal of Epidemiology 2019;48(6):1783–94. doi: 10.1093/ije/dyz129

8. Huttunen R, Syrjänen J. Obesity and the risk and outcome of infection. International Journal of Obesity 2013;37(3):333–40. doi: 10.1038/ijo.2012.62

9. Winter-Jensen M, Afzal S, Jess T, et al. Body mass index and risk of infections: a Mendelian randomization study of 101,447 individuals. Eur J Epidemiol 2020;35:347–54. doi: 10.1007/s10654-020-00630-7 [published Online First: 2020/04/21]

10. Kaspersen KA, Pedersen OB, Petersen MS, et al. Obesity and Risk of Infection: Results from the Danish Blood Donor Study. Epidemiology 2015;26(4)

11. Paulsen J, Askim Å, Mohus RM, et al. Associations of obesity and lifestyle with the risk and mortality of bloodstream infection in a general population: a 15-year follow-up of 64?027 individuals in the HUNT Study. Int J Epidemiol 2017;46(5):1573–81. doi: 10.1093/ije/dyx091 [published Online First: 2017/06/24]

12. Guan WJ, Ni ZY, Hu Y, et al. Clinical Characteristics of Coronavirus Disease 2019 in China. N Engl J Med 2020;382(18):1708–20. doi: 10.1056/NEJMoa2002032

13. Docherty AB, Harrison EM, Green CA, et al. Features of 20 133 UK patients in hospital with covid-19 using the ISARIC WHO Clinical Characterisation Protocol: prospective observational cohort study. BMJ 2020;369 doi: 10.1136/bmj.m1985

14. Liu W, Tao Z-W, Lei W, et al. Analysis of factors associated with disease outcomes in hospitalized patients with 2019 novel coronavirus disease. Chinese Medical Journal 9000;Publish Ahead of Print

15. Simonnet A, Chetboun M, Poissy J, et al. High prevalence of obesity in severe acute respiratory syndrome coronavirus-2 (SARS-CoV-2) requiring invasive mechanical ventilation. Obesity 2020 doi: 10.1002/oby.22831

16. Williamson E, Walker AJ, Bhaskaran KJ, et al. OpenSAFELY: factors associated with COVID-19-related hospital death in the linked electronic health records of 17 million adult NHS patients. medRxiv 2020:2020.05.06.20092999. doi: 10.1101/2020.05.06.20092999

17. Ng PY, Eikermann M. The obesity conundrum in sepsis. BMC Anesthesiol 2017;17(1):147–47. doi: 10.1186/s12871-017-0434-z

18. Niskanen L, Laaksonen DE, Nyyssönen K, et al. Inflammation, Abdominal Obesity, and Smoking as Predictors of Hypertension. Hypertension 2004;44(6):859–65. doi: doi:10.1161/01.HYP.0000146691.51307.84

19. Lawlor DA, Harbord RM, Sterne JAC, et al. Mendelian randomization: Using genes as instruments for making causal inferences in epidemiology. Statistics in Medicine 2008;27(8):1133–63. doi: 10.1002/sim.3034

20. Walley KR, Boyd JH, Kong HJ, et al. Low Low-Density Lipoprotein Levels Are Associated With, But Do Not Causally Contribute to, Increased Mortality in Sepsis*. Critical Care Medicine 2019;47(3)

21. Trinder M, Walley KR, Boyd JH, et al. Causal Inference for Genetically Determined Levels of High-Density Lipoprotein Cholesterol and Risk of Infectious Disease. Arteriosclerosis, Thrombosis, and Vascular Biology 2020;40(1):267–78. doi: doi:10.1161/ATVBAHA.119.313381

22. Ellinghaus D, Degenhardt F, Bujanda L, et al. Genomewide Association Study of Severe Covid-19 with Respiratory Failure. New England Journal of Medicine 2020 doi: 10.1056/NEJMoa2020283

23. Hemani G, Zheng J, Elsworth B, et al. The MR-Base platform supports systematic causal inference across the human phenome. eLife 2018;7 doi: 10.7554/eLife.34408

24. Willer CJ, Schmidt EM, Sengupta S, et al. Discovery and refinement of loci associated with lipid levels. Nat Genet 2013;45(11):1274–83. doi: 10.1038/ng.2797

25. Sudlow C, Gallacher J, Allen N, et al. UK biobank: an open access resource for identifying the causes of a wide range of complex diseases of middle and old age. PLoS Med 2015;12(3):e1001779–e79. doi: 10.1371/journal.pmed.1001779

26. Bycroft C, Freeman C, Petkova D, et al. The UK Biobank resource with deep phenotyping and genomic data. Nature 2018;562(7726):203–09. doi: 10.1038/s41586-018-0579-z

27. Mitchell R, Elsworth B, Mitchell R, et al. MRC IEU UK Biobank GWAS pipeline version 2. University of Bristol Data Repository 2019

28. Krokstad S, Langhammer A, Hveem K, et al. Cohort Profile: the HUNT Study, Norway. Int J Epidemiol 2013;42(4):968–77. doi: 10.1093/ije/dys095 [published Online First: 2012/08/11]

29. Ferreira MA, Vonk JM, Baurecht H, et al. Shared genetic origin of asthma, hay fever and eczema elucidates allergic disease biology. Nature genetics 2017;49(12):1752–57. doi: 10.1038/ng.3985 [published Online First: 2017/10/30]

30. Zhou W, Nielsen JB, Fritsche LG, et al. Efficiently controlling for case-control imbalance and sample relatedness in large-scale genetic association studies. Nature Genetics 2018;50(9):1335–41. doi: 10.1038/s41588-018-0184-y

31. Wang C, Zhan X, Liang L, et al. Improved ancestry estimation for both genotyping and sequencing data using projection procrustes analysis and genotype imputation. American journal of human genetics 2015;96(6):926–37. doi: 10.1016/j.ajhg.2015.04.018 [published Online First: 2015/05/28]

32. Wang C, Zhan X, Bragg-Gresham J, et al. Ancestry estimation and control of population stratification for sequence-based association studies. Nature genetics 2014;46(4):409–15. doi: 10.1038/ng.2924 [published Online First: 2014/03/16]

33. Burgess S, Butterworth A, Thompson SG. Mendelian randomization analysis with multiple genetic variants using summarized data. Genet Epidemiol 2013;37(7):658–65. doi: 10.1002/gepi.21758 [published Online First: 2013/09/20]

34. Burgess S, Bowden J, Fall T, et al. Sensitivity Analyses for Robust Causal Inference from Mendelian Randomization Analyses with Multiple Genetic Variants. Epidemiology (Cambridge, Mass) 2017;28(1):30–42. doi: 10.1097/EDE.0000000000000559

35. Bowden J, Davey Smith G, Haycock PC, et al. Consistent Estimation in Mendelian Randomization with Some Invalid Instruments Using a Weighted Median Estimator. Genet Epidemiol 2016;40(4):304–14. doi: 10.1002/gepi.21965 [published Online First: 2016/04/07]

36. Hartwig FP, Davey Smith G, Bowden J. Robust inference in summary data Mendelian randomization via the zero modal pleiotropy assumption. International journal of epidemiology 2017;46(6):1985–98. doi: 10.1093/ije/dyx102

37. Burgess S, Foley CN, Allara E, et al. A robust and efficient method for Mendelian randomization with hundreds of genetic variants. Nature Communications 2020;11(1):376. doi: 10.1038/s41467-019-14156-4

38. Bowden J, Davey Smith G, Burgess S. Mendelian randomization with invalid instruments: effect estimation and bias detection through Egger regression. International journal of epidemiology 2015;44(2):512–25. doi: 10.1093/ije/dyv080 [published Online First: 2015/06/06]

39. Sanderson E, Davey Smith G, Windmeijer F, et al. An examination of multivariable Mendelian randomization in the single-sample and two-sample summary data settings. Int J Epidemiol 2018:48(3):713–27. doi: 10.1093/ije/dyy262

40. Burgess S, Thompson SG. Multivariable Mendelian randomization: the use of pleiotropic genetic variants to estimate causal effects. Am J Epidemiol 2015;181(4):251–60. doi: 10.1093/aje/kwu283

41. Burgess S, Dudbridge F, Thompson SG. Re: “Multivariable Mendelian randomization: the use of pleiotropic genetic variants to estimate causal effects”. Am J Epidemiol 2015;181(4):290–1. doi: 10.1093/aje/kwv017

42. Yavorska OO, Burgess S. MendelianRandomization: an R package for performing Mendelian randomization analyses using summarized data. International journal of epidemiology 2017;46(6):1734–39. doi: 10.1093/ije/dyx034

43. Davey Smith G, Davies NM, Dimou N, et al. STROBE-MR: Guidelines for strengthening the reporting of Mendelian randomization studies. https://doi.org/10.7287/peerj.preprints.27857v1. PeerJ Preprints 2019;7:pe27857v1

44. Pépin JL, Timsit JF, Tamisier R, et al. Prevention and care of respiratory failure in obese patients. The Lancet Respiratory Medicine 2016;4(5):407–18. doi: 10.1016/S2213-2600(16)00054-0

45. Fröhlich M, Sund M, Löwel H, et al. Independent association of various smoking characteristics with markers of systemic inflammation in men: Results from a representative sample of the general population (MONICA Augsburg Survey 1994/95). European Heart Journal 2003;24(14):1365–72. doi: 10.1016/s0195-668x(03)00260-4

46. Kuschner W, D’Alessandro A, Wong H, et al. Dose-dependent cigarette smoking-related inflammatory responses in healthy adults. European Respiratory Journal 1996;9(10):1989–94.

47. Laing AG, Lorenc A, Del Molino Del Barrio I, et al. A consensus Covid-19 immune signature combines immuno-protection with discrete sepsis-like traits associated with poor prognosis. medRxiv 2020:2020.06.08.20125112. doi: 10.1101/2020.06.08.20125112

48. Xu Z, Shi L, Wang Y, et al. Pathological findings of COVID-19 associated with acute respiratory distress syndrome. The Lancet Respiratory Medicine 2020;8(4):420–22. doi: 10.1016/S2213-2600(20)30076-X

49. Shen B, Yi X, Sun Y, et al. Proteomic and Metabolomic Characterization of COVID-19 Patient Sera. Cell 2020 doi: https://doi.org/10.1016/j.cell.2020.05.032

50. Roschewski M, Lionakis MS, Sharman JP, et al. Inhibition of Bruton tyrosine kinase in patients with severe COVID-19. Science Immunology 2020;5(48):eabd0110. doi: 10.1126/sciimmunol.abd0110

51. Wadud N, Ahmed N, Mannu Shergil M, et al. Improved survival outcome in SARs-CoV-2 (COVID-19) Acute Respiratory Distress Syndrome patients with Tocilizumab administration. medRxiv 2020:2020.05.13.20100081. doi: 10.1101/2020.05.13.20100081

52. RECOVERY (Randomised Evaluation of COVid-19 thERapY) trial. Low-cost dexamethasone reduces death by up to one third in hospitalised patients with severe respiratory complications of COVID-19. https://www.recoverytrial.net/files/recovery_dexamethasone_statement_160620_final.pdf: Oxford University News Release, 2020.

53. Russell JAaT, Mark and Lee, Terry and Brunham, Liam R. and Hsu, Joe and Boyd, John and Singer, Joel and Walley, Keith R.,. Diabetes Increases Risk of Sepsis, But Not Mortality, Inflammatory or Lipid Responses. Lancet Preprint 2019;Available http://dx.doi.org/10.2139/ssrn.3387518

54. Afshin A, Collaborators. TGO. Health Effects of Overweight and Obesity in 195 Countries over 25 Years. New England Journal of Medicine 2017;377(1):13–27. doi: 10.1056/NEJMoa1614362

55. Ward ZJ, Bleich SN, Cradock AL, et al. Projected U.S. State-Level Prevalence of Adult Obesity and Severe Obesity. New England Journal of Medicine 2019;381(25):2440–50. doi: 10.1056/NEJMsa1909301

56. Keaver L, Xu B, Jaccard A, et al. Morbid obesity in the UK: A modelling projection study to 2035. Scand J Public Health 2018:1403494818794814. doi: 10.1177/1403494818794814 [published Online First: 2018/08/31]

57. Baskaran V, Murray RL, Hunter A, et al. Effect of tobacco smoking on the risk of developing community acquired pneumonia: A systematic review and meta-analysis. PloS one 2019;14(7):e0220204–e04. doi: 10.1371/journal.pone.0220204

58. Lawrence H, Hunter A, Murray R, et al. Cigarette smoking and the occurrence of influenza - Systematic review. Journal of Infection 2019;79(5):401–06. doi: 10.1016/j.jinf.2019.08.014

59. Ng M, Freeman MK, Fleming TD, et al. Smoking prevalence and cigarette consumption in 187 countries, 1980-2012. JAMA 2014;311(2):183–92. doi: 10.1001/jama.2013.284692 [published Online First: 2014/01/09]

60. Cornish D, Brookman A, Horton M, et al. Adult smoking habits in the UK: 2018. https://www.ons.gov.uk/peoplepopulationandcommunity/healthandsocialcare/healthandlifeexpectancies/bulletins/adultsmokinghabitsingreatbritain/2018: Office for National Statistics, 2018.

61. Lasser K, Boyd JW, Woolhandler S, et al. Smoking and mental illness: A population-based prevalence study. JAMA 2000;284(20):2606–10. doi: 10.1001/jama.284.20.2606 [published Online First: 2000/11/22]

62. Hickman NJ, 3rd, Delucchi KL, Prochaska JJ. A population-based examination of cigarette smoking and mental illness in Black Americans. Nicotine Tob Res 2010;12(11):1125–32. doi: 10.1093/ntr/ntq160 [published Online First: 2010/09/22]

63. Annamalai A, Kosir U, Tek C. Prevalence of obesity and diabetes in patients with schizophrenia. World J Diabetes 2017;8(8):390–96. doi: 10.4239/wjd.v8.i8.390

64. Pankiewicz-Dulacz M, Stenager E, Chen M, et al. Incidence Rates and Risk of Hospital Registered Infections among Schizophrenia Patients before and after Onset of Illness: A Population-Based Nationwide Register Study. J Clin Med 2018;7(12):485. doi: 10.3390/jcm7120485

65. Slob EAW, Burgess S. A comparison of robust Mendelian randomization methods using summary data. Genet Epidemiol 2020;44(4):313–29. doi: 10.1002/gepi.22295

66. Burgess S, Labrecque JA. Mendelian randomization with a binary exposure variable: interpretation and presentation of causal estimates. European Journal of Epidemiology 2018;33(10):947–52. doi: 10.1007/s10654-018-0424-6

67. Critchley JA, Carey IM, Harris T, et al. Glycemic Control and Risk of Infections Among People With Type 1 or Type 2 Diabetes in a Large Primary Care Cohort Study. Diabetes Care 2018;41(10):2127–35. doi: 10.2337/dc18-0287

68. Pulit SL, Stoneman C, Morris AP, et al. Meta-analysis of genome-wide association studies for body fat distribution in 694 649 individuals of European ancestry. Hum Mol Genet 2019;28(1):166–74. doi: 10.1093/hmg/ddy327

69. Wootton RE, Richmond RC, Stuijfzand BG, et al. Evidence for causal effects of lifetime smoking on risk for depression and schizophrenia: a Mendelian randomisation study. Psychol Med 2019:1–9. doi: 10.1017/S0033291719002678

70. Carter AR, Gill D, Davies NM, et al. Understanding the consequences of education inequality on cardiovascular disease: mendelian randomisation study. BMJ 2019;365:1855. doi: 10.1136/bmj.l1855

71. Mahajan A, Taliun D, Thurner M, et al. Fine-mapping type 2 diabetes loci to single-variant resolution using high-density imputation and islet-specific epigenome maps. Nat Genet 2018;50(11):1505–13. doi: 10.1038/s41588-018-0241-6

